# Impact of the COVID-19 pandemic on surgical procedures in Brazil: a descriptive study

**DOI:** 10.1101/2021.03.17.21253801

**Authors:** Bárbara Okabaiasse Luizeti, Victor Augusto Santos Perli, Gabriel Gonçalves da Costa, Igor da Conceição Eckert, Aluisio Marino Roma, Karina Miura da Costa

## Abstract

**Background:** The COVID-19 pandemic has deeply affected medical practice, and changes in healthcare activities were needed to minimize the overload and avoid healthcare systems collapse. The aim of this study was to evaluate the impact of the pandemic on surgical procedures in Brazil.

**Materials and Methods:** We conducted a descriptive study of the number of hospitalizations for surgical procedures in Brazil from 2016 to 2020. Data were collected from the Brazilian Department of Informatics of the Unified Health System (DATASUS). Analyzes were performed according to the type of procedure, geographical region, subgroups of surgical procedures, and the number of surgeries from 2020 were compared with the average from 2016 to 2019.

**Results:** There were 4,009,116 hospitalizations for surgical procedures in the Brazilian Public Health System in 2020. When comparing it to the average of hospitalizations from 2016-2019, there was a decrease of 14.88% [95%IC (14.82-14.93)]. Decrease rates were 34.82% [95%IC (34.73-34.90)] for elective procedures and 1.11% [95%IC (1.07-1.13)] for urgent procedures. Decrease rates were similar in all the five regions of the country (average 14.17%). Surgical procedure subgroups with the highest decrease rates were endocrine gland surgery (48.03%), breast surgery (40.68%), oral and maxillofacial surgery (37.03%), surgery of the upper airways, face, head and neck (36.06%), and minor surgeries and surgeries of skin, subcutaneous tissue and mucosa (33.16%). Conclusion: The overload of healthcare facilities has demanded a reduction of non-urgent activities to prevent a collapse of healthcare systems, resulting in a decrease in elective surgeries. Recommendations about the performance of surgical procedures were made, and continuous refinements of these recommendations are encouraged.

## 1. Introduction

Coronavirus Disease 2019 (COVID-19) was first reported in late December 2019 in Wuhan, Hubei Province of China [1,2]. The disease spread quickly: by the end of January 2020, it was stated as an international public health emergency, and, in March, the World Health Organization (WHO) declared COVID-19 as a pandemic [3,4].

Since the beginning of the outbreak, there was concern about the impact on healthcare systems. Due to the risk of nosocomial transmission, possible lack of personal protective equipment (PPE), overflow of healthcare facilities, and limited medical resources [5,6], there was a disruption in delivering surgical care to millions of patients [7-9].

The relation between surgery and the outbreak is complex: the risk of exposure and nosocomial infection is higher in the context of a surgical procedure; postoperative care may need hospitalization and intensive care, thus reducing the availability of these supports to COVID-19 patients. Other situations affecting surgical practice were also seen, such as the shortage of medications and blood components [7,8].

In this context, many countries canceled elective procedures to redirect surgical resources and workforce to COVID-19 patients [9-11]. Furthermore, considering that the healthcare professionals could be at high risk of exposure to the virus during surgical procedures [12], specific institutional protocols were developed for operating rooms depending on the type of surgery [13,14].

The aim of this study was to analyze the impact of the COVID-10 pandemic on surgical procedures in Brazil.

## 2. Materials and Methods

This was a descriptive study, based on publicly available data from the Brazilian Department of Informatics of the Unified Health System (DATASUS), which includes only data from the Brazilian public health system, collected in the public health information tab – TABNET.

Data from 2016 to 2020 were collected according to the procedure and Brazil’s geographical regions. Brazil is divided into five regions encompassing 27 federative units: North (seven federative units); Northeast (nine federative units); Midwest (four federative units); Southeast (four federative units); and South (three federative units) [15].

Data were also collected according to the type of procedure, which was classified in TABNET as “elective procedure”, “urgent procedure”, “accident in the workplace or at the duty of a company”, “accident on the way to work”, “other types of traffic accidents” and “other types of injury or poisoning by chemical or physical agents”. The last four were computed in a single group called “other procedures”.

Historical time series charts were performed by month and by year for the variables mentioned above. For 2020, hospitalizations for total surgical procedures were listed per month and per region. Analyses were performed by type of surgical procedure and by region, comparing the average of hospitalizations rates from 2016 to 2019 with the hospitalization rates in 2020.

Surgical procedures subgroups were analyzed, comparing the averages of hospitalizations from 2016 to 2019 with hospitalizations in 2020 to determine the subgroups most affected by the COVID-19 pandemic.

Analyses were performed in software R for statistical computing, version 4.0.3 (R Core Team, Vienna, Austria). This article is compliant with the STROCSS criteria (Strengthening the reporting of cohort studies in surgery).

## 3. Results

The number of hospitalizations for surgical procedures from 2016 to 2020 is shown in Table 1 and Figure 1, with temporal distribution of total, elective and urgent surgical procedures. There was an increase in surgeries from 2016 to 2019, and a reduction in 2020. When comparing the average of hospitalizations from 2016-2019 and the hospitalizations in 2020, there was a 14.88% reduction [95%IC (14.82-14.93)] in total surgical procedures (Table 1). Elective procedures showed a greater decrease, with 34.82% [95%IC (34.73-34.90)] less elective procedures and 1.11% [95%IC (1.07-1.13)] less urgent procedures in 2020.

**Table 1.** Number of hospitalizations for surgical procedures in Brazil between 2016 and 2020, according to the type of procedure.

| Procedure | 2016 | 2017 | 2018 | 2019 | 2020 |
| --- | --- | --- | --- | --- | --- |
| **Elective surgeries** | 1,726,279 | 1,790,425 | 1,984,641 | 2,089,807 | 1,237,056 |
| **Urgency surgeries** | 2,644,140 | 2,716,817 | 2,774,911 | 2,844,255 | 2,714,709 |
| **Other procedures** | 73,988 | 66,823 | 64,503 | 62,901 | 57,351 |
| **Total surgeries** | 4,444,407 | 4,574,065 | 4,824,055 | 4,996,963 | 4,009,116 |

**Figure 1.**
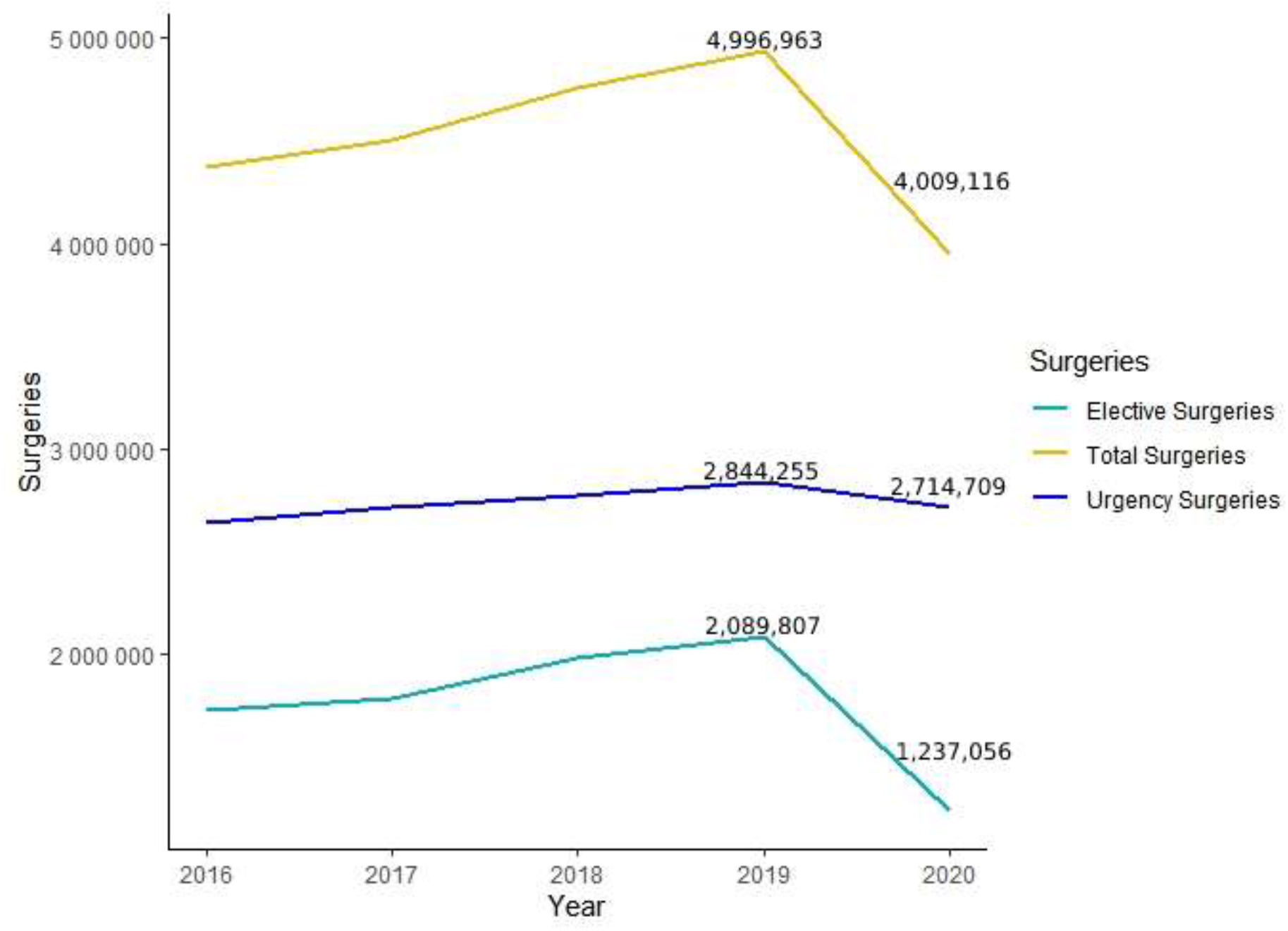
Temporal evolution of the number of elective, urgent and total surgical procedures in Brazil between 2016 and 2020.

The number of surgical hospitalizations in each region between 2016 and 2020 is shown in Table 2. Comparing hospitalizations from 2016 to 2019 and in 2020, there was a 15.81% reduction in the Southeast, 14.96% in the Northeast, 14.68% in the North, 14.37% in the South, and 11.04% in the Midwest (average reduction of 14,17%).

**Table 2.** Number of hospitalizations for surgical procedures in each of the five Brazilian regions, 2016-2020.

| Procedure | 2016 | 2017 | 2018 | 2019 | 2020 |
| --- | --- | --- | --- | --- | --- |
| **North** | 325,975 | 342,398 | 360,933 | 366,708 | 297,765 |
| **Northeast** | 1,160,769 | 1,189,679 | 1,286,596 | 1,321,188 | 1,054,120 |
| **Midwest** | 343,797 | 348,354 | 357,152 | 377,746 | 317,390 |
| **Southeast** | 1,801,439 | 1,855,185 | 1,944,753 | 2,020,534 | 1,604,257 |
| **South** | 812,427 | 838,449 | 874,621 | 910,787 | 735,584 |
| **Total** | 4,444,407 | 4,574,065 | 4,824,055 | 4,996,963 | 4,009,116 |

The number of hospitalizations according to the surgical procedures subgroup can be seen in Table 3. Endocrine gland surgery, breast surgery, oral/maxillofacial surgeries, surgery of the upper airways/face/head/neck, and minor surgeries/surgeries of skin/subcutaneous tissue/mucosa were the five subgroups with the highest decrease rates. The lowest decreases were found in reconstructive, osteomuscular, thoracic, oncology and other surgery subgroups. Obstetric surgery was the only subgroup with an increased rate (0.48%).

**Table 3.** Number of hospitalizations according to the subgroups of surgical procedures between 2016 and 2020 in Brazil.

| Surgical procedures subgroups | Hospitalizations from 2016-2019 (average) | Hospitalizations in 2020 | Ratio (%) |
| --- | --- | --- | --- |
| Endocrine gland surgery | 12,253 | 6,368 | -48.03% |
| Breast surgery | 34,824.75 | 20,660 | -40.68% |
| Oral and maxillofacial surgery | 13,273.75 | 8,322 | -37.3% |
| Surgery of the upper airways, face, head and neck | 132,504.5 | 84,719 | -36.06% |
| Minor surgeries and surgeries of skin, subcutaneous tissue and mucosa | 118,676 | 79,322 | -33.16% |
| Surgery of the genitourinary system | 501,674.5 | 338,983 | -32.43% |
| Vision apparatus surgery | 108,136.5 | 73,779 | -31.77% |
| Surgery of the digestive system, attached organs and abdominal wall | 768,490.5 | 536,729 | -30.16% |
| Circulatory system surgery | 292,014 | 234,947 | -19.54% |
| Central and peripheral nervous system surgery | 88,425.75 | 72,765 | -17.71% |
| Reconstructive surgery | 57,821.25 | 47,587 | -17.70% |
| Surgery of the osteomuscular system | 768,984.25 | 701,006 | -8.84% |
| Thoracic surgery | 58,799.5 | 55,258 | -6.02% |
| Oncological surgery | 146,922.5 | 138,701 | -5.60% |
| Other surgeries | 557,951.5 | 556,027 | -0.35% |
| Obstetric surgery | 1,048,853.75 | 1,053,943 | +0.48% |
| Total | 4,709,872.5 | 4,009,116 | -14.88% |

Figure 2 shows the number of hospitalizations per month in 2020. Regarding the 4,009,116 surgical procedures performed, there was a reduction from the beginning to the middle of the year: January (403,829 procedures), February (396,118 procedures [-1.90%]), March (380,610 procedures [-3.91%]), April (285.924 procedures [-24.87%]), May (285.071 procedures [-0.29%]) and June (276,959 procedures [-2.84%]). There was an average decrease rate of 6.77% in this period, with the highest decrease observed from March to April. From July to November, there was an increase in the number of surgeries: July (299,058 procedures), August (310,016 procedures [+3.66%]), September (329,551 procedures [+6.30%]), October (360,078 procedures [+9.26%]) and November (361,027 procedures [+0.26%]), with an average increase rate of 5.49% in this period. In December (320,875 procedures), there is a new decrease rate of 11.12% compared to November.

**Figure 2.**
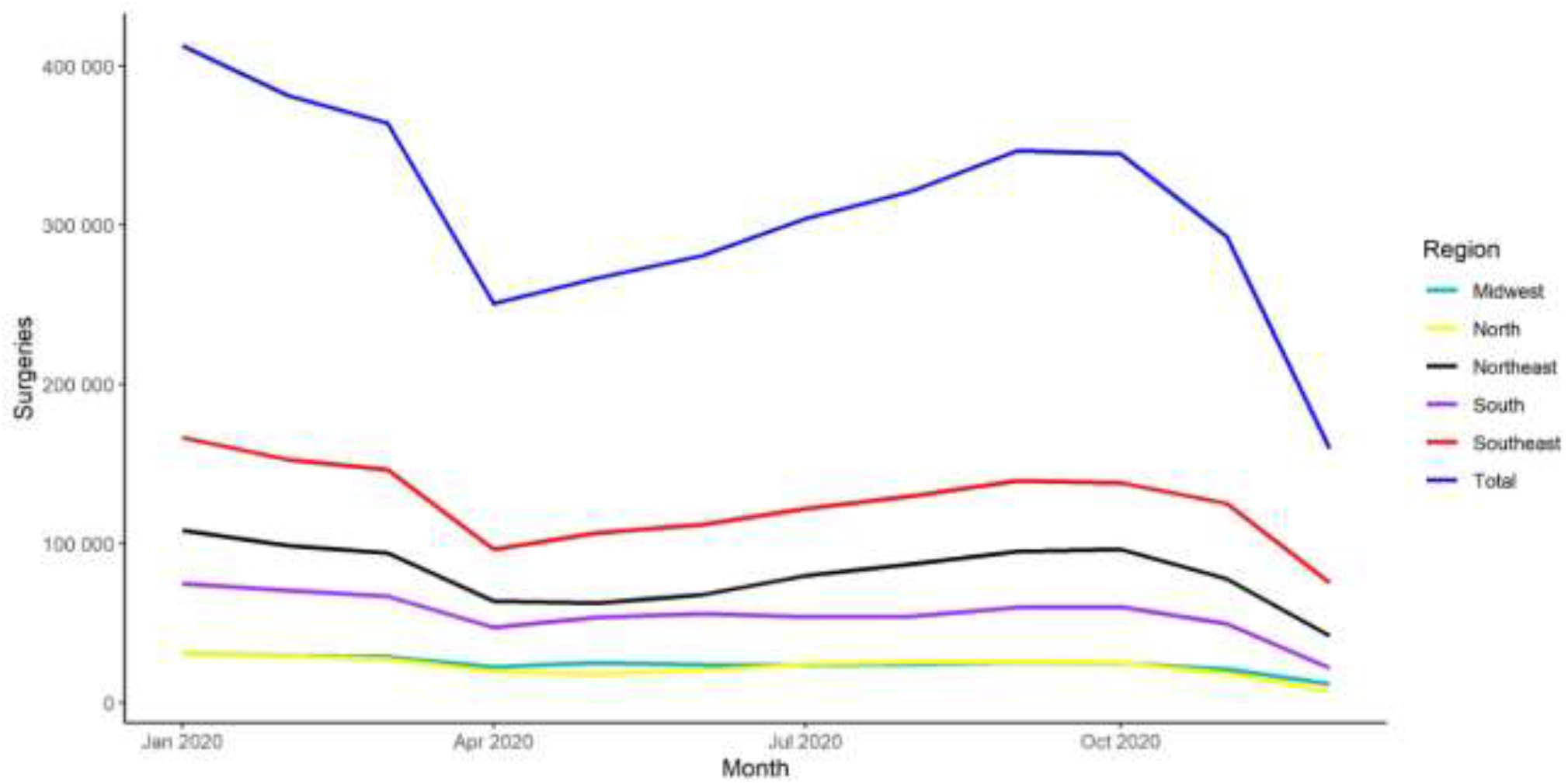
Number hospitalizations for surgical procedures in 2020, per month.

Figure 2 also shows the number of hospitalizations per month in 2020 according to the country’s regions. When comparing the hospitalization rates in January and December, there were decreased rates of 36.58% in North, 22.06% in South, 19.95% in Southeast, 19.25% in Midwest, and 16.49% Northeast. As well as in the overall analysis, the highest reductions occurred in April and December. In April, there were decreased rates of 27.08% in Southeast, 24.88% in Midwest, 24.74% in South, 23.74% in Northeast and 21.2% in North. In December, decrease rates were 26.73% in North, 12.73% in Northeast, 10.31% in South, 9.15% in Southeast and 1.82% in Midwest.

According to the National Register of Health Establishments of the Ministry of Health of Brazil, in December 2020, 332,192 health establishments were registered with DATASUS, of which 17,283 in the North, 67,442 in the Northeast, 146,558 in the Southeast, 73,857 in the South and 27,052 in the Midwest. In December 2019, 345,017 were registered with DATASUS, with 17,480 in the North, 68,297 in the Northeast, 162,642 in the Southeast, 70,998 in the South and 25,600 in the Midwest. In December 2018, 331,058 were registered with DATASUS, with 16,958 in the North, 66,343 in the Northeast, 155,229 in the Southeast, 68,105 in the South and 24,423 in the Midwest. In December 2017, 312,783 were registered with DATASUS, with 15,775 in the North, 63,164 in the Northeast, 146,432 in the Southeast, 64,450 in the South and 22,962 in the Midwest. In December 2016, 298,173 were registered with DATASUS, 15,006 in the North, 60,177 in the Northeast, 137,751 in the Southeast, 61,200 in the South and 24,039 in the Midwest [15].

## 4. Discussion

The decision to perform surgeries in the face of the pandemic requires a thorough evaluation to establish priorities and define precisely in which cases benefits overcome risks, and surgery is genuinely needed [9,16]. This was a great challenge for surgeons in 2020, since avoiding unnecessary activities in healthcare facilities, thus canceling or postponing all non-urgent procedures, was widely stated as an important measure to prevent overload and enable reorganization of the healthcare systems worldwide [9-11]. The Brazilian public health system adhered to these recommendations, as shown by the decrease of elective surgeries in 2020 (34.82%) compared to the decline in urgent procedures (1.1%).

When analyzing monthly surgical procedures in 2020, there was a remarkably low number of surgeries between April and June, the months following the country’s first reported cases, resuming growth from July onwards. Indeed, regulatory measures must be more restrictive at the beginning of an outbreak, when healthcare systems are unprepared to deal with a possible overload, so time is needed until more health resources can be acquired [9,10]. There was a new reduction rate in December when compared to the previous months. In the past years (2017-2019), there was a seasonal decrease in hospitalizations for surgical procedures in November and December [17]. Still, the reduction in 2020 could also be attributed to the increase in COVID-19 cases that took part in Brazil in November [18].

Even though Brazil has considerable differences in public administration, heterogeneous cultural and economic conditions across different regions, data shows similar reduction rates in hospitalizations when comparing the rates in 2020 with the previous years, with an average reduction of 18.96%. However, in the 2020 monthly comparison, there was a higher decrease rate in the North in December when compared to the other regions. These reductions might have been the result of the regional healthcare system collapse in December [19].

Surgical care that is not time-critical can be delayed to a later date when the pandemic subsides. However, certain procedures must be performed even during a pandemic, such as cancer treatment and urgent surgeries, as these are considered life-saving procedures [9]. This could explain why oncology and thoracic surgeries suffered minor reductions in the ranking, while endocrine gland surgery was the subgroup with the highest decrease rates (48.03%), probably due to the possibility of safe deferral of these procedures [16,20,21].

Regarding plastic surgery, guidelines support the deferral of breast reconstruction and revision procedures [22], which might explain the high decrease rates in breast surgery (40.68%). Other studies have found that plastic surgery procedures most impacted by the pandemic were superficial skin lesions and subcutaneous mass procedures [23], which corroborates our findings of high decrease rates in minor surgeries and surgeries of skin, subcutaneous tissue and mucosa (33.16%).

Other subgroups with the highest decrease rates were oral and maxillofacial (37.3%) and upper airways, head and neck (36.06%) surgeries. These procedures were of particular concern due to the higher risk of generating aerosols. Guidelines recommend these surgeries should only be performed in life-threatening conditions with great precautions due to the possibility of COVID-19 infection [22,24].

There were also important decrease rates among surgeries of the digestive system and abdominal wall (30.16%), which may also reflect recommendations and alternative management of digestive emergency conditions performed during the pandemic. Guidelines support that only life-threatening gastrointestinal conditions should undergo surgery, and there were recommendations for clinical treatment in conditions such as uncomplicated appendicitis and cholecystitis [17,23], situations in which conservative management appears to be effective and reduce the length of hospital stay [25].

Cancer surgery was one of the challenges faced in the context of the pandemic. Since both cancer and surgical patients are at high risk of COVID-19 infection and complications of the disease [26-28], there was a concern involving the exposure of these patients to the hospital environment, and most guidelines supported that surgeries should be delayed whenever possible. On the other hand, there were concerns about the risks of postponing oncological procedures and the exposure of patients undergoing conservative cancer treatment. The recommendation was that decisions should be made based on the clinical condition and the pandemic’s impact on the local healthcare system [22,27,28]. This study found only a 5.6% reduction in general oncological surgeries, which suggests that Brazil had a trend in choosing for operation in most cancer cases.

Hospitalization rates for obstetric surgery were slightly higher in 2020 than the average from 2016 to 2019 (0.48% increase). Brazil is one of the countries with the highest cesareans rates, which consisted of 56.3% of all deliveries in 2019 [29]. Since most guidelines support that maternal COVID-19 infection should not influence the type of delivery [30], it should be safe to assume that the pandemic did not significantly impact this group of procedures.

COVID-19 has had a devastating impact on patients, the economy and healthcare systems. There is hope that cases will begin to decline soon and, as the COVID-19 vaccine is distributed, hospitals must develop protocols to resume elective surgeries safely. This will be the next challenge for the surgical community. Moving forward requires not only creativity but also structural changes to patient workflows [31-33].

Regarding the long-term effects on surgical practice, this pandemic should raise awareness regarding protective measures to both surgical staff and patients, and the protocols’ changes made to reduce the risk of contamination during surgical procedures should be added to operating theaters routines.

This study has limitations: the data refer only to the Brazilian public health system because data from the private health system are not available, which might lead to an overestimation of the decrease rates. As secondary data, the veracity of the information provided depends on proper registration.

Although regression analyses that could help explain the differences in the number of surgeries between subgroups and regions were not performed, some justifications for the temporal trends of reduction in surgical procedures are speculated, such as the correlation with decrees and laws of state and municipal governments that imposed the postponement of elective surgeries.

## 5. Conclusion

COVID-19 pandemic has promoted changes in the surgical practice in Brazil. The need for minimizing the overload in healthcare facilities has demanded reductions in healthcare activities and avoidance of non-urgent procedures, as seen by the notable decline in elective surgeries. Lower numbers of procedures were seen in April, July and December of 2020, periods of an increasing number of cases in the country.

Continuous refinements of safety recommendations are encouraged because of evolving circumstances. The new protocols developed during the pandemic should be implemented and improved for patients and staff safety in the long term.

## Data Availability

To foster transparency, we state the availability of our data for access.

https://cnes.datasus.gov.br

http://tabnet.datasus.gov.br/cgi/deftohtm.exe?sih/cnv/qiuf.def

http://tabnet.datasus.gov.br/cgi/deftohtm.exe?sih/cnv/qiuf.def

## Declarations of interest

None.

## Funding

This research did not receive any specific grant from funding agencies in the public, commercial, or not-for-profit sectors.

## CRediT author statement

Bárbara Okabaiasse Luizeti: Conceptualization; Methodology; Investigation; Writing – Original Draft; Project administration. Victor Augusto Santos Perli: Methodology; Investigation; Resources; Writing – Original Draft. Gabriel Gonçalves da Costa: Software; Formal analysis; Data Curation; Writing – Review & Editing. Igor da Conceição Eckert: Validation; Writing – Review & Editing. Aluisio Marino Roma: Validation; Visualization. Karina Miura da Costa: Writing – Original Draft; Supervision.

## Notes

### Competing Interest Statement

The authors have declared no competing interest.

### Author Declarations

To foster transparency, we state the availability of our data for access.

